# Neighborhood socioeconomic status associated with post-stroke cognitive impairment: a retrospective cohort study

**DOI:** 10.64898/2026.06.09.26355320

**Authors:** Matthew Siegel, Laura Corlin, James Miller, Kathryn Cote, Lester Y Leung

**Affiliations:** Tufts University School of Medicine, Boston; Department of Public Health and Community Medicine, Tufts University School of Medicine, Boston; Department of Community Health, Tufts University School of Arts and Sciences, Medford; Department of Civil and Environmental Engineering, Tufts University School of Engineering, Medford; Montefiore Medical Center, Bronx; Department of Vascular Neurology, Tufts Medical Center, Boston

**Keywords:** stroke, cognitive impairment, social determinants, inequality

## Abstract

**Background:** Late complications after stroke (LCAS), including cognitive symptoms, impact quality of life and recovery. It is not known if neighborhood-level measures of socioeconomic status (SES) influence LCAS. This study assessed associations between SES measures, including neighborhood income inequality (Gini) and area deprivation index (ADI), and cognitive symptoms after acute ischemic stroke (AIS) in a hospital leveraging active surveillance of LCAS.

**Methods:** This retrospective cohort study included 512 patients hospitalized with AIS at Tufts Medical Center with subsequent follow-up (between zero and three months or between three and twelve months) in the Stroke Clinic from 1/1/2018 - 12/31/2022. Using ZIP code data, patients were characterized as low Gini (low inequality) and high ADI (high deprivation) (Gini <= 0.4302, ADI >= 5) by state medians. These variables were combined, indicating patients who were living in both a low Gini and high ADI neighborhood to evaluate the effects of living in a homogeneously deprived area. There were 206 and 281 patients in the low Gini and high ADI groups respectively. 140 patients lived in a low Gini and high ADI neighborhood. The multivariable logistic analysis assessed the likelihood of cognitive symptoms, adjusting for age, race, ethnicity, sex, NIH Stroke Scale (NIHSS), thrombolysis, active LCAS surveillance, poverty, and ADI-Gini combination.

**Results:** There were no associations between high ADI (OR: 1.03, 95% CI: 0.67 – 1.57) or low Gini (OR: 1.74, 95% CI: 0.98 – 3.07) alone and cognitive symptoms after AIS. However, the combined variable demonstrated increased likelihood of cognitive symptoms in the high ADI-low Gini group (OR: 1.82, 95% CI: 1.08 – 3.06).

**Conclusions:** This study suggests that individuals living in homogeneously deprived neighborhoods report higher likelihood of cognitive symptoms after AIS. Further studies with increased power are needed to investigate the underlying causes of these disparities and to develop interventions to reduce these complications.

## Introduction

While social determinants of health are known to be associated with frequency of acute treatment and functional outcomes, it is not certain if these social determinants are linked to post-stroke complications such as cognitive impairment.^1,2^ Late complications after stroke (LCAS) are common and an area of emerging research.^3–5^ LCAS encompass a wide range of conditions including some that are more visible (e.g., spasticity) and others that are referred to by patients as “invisible disabilities” (including cognitive impairment).^5^ There have been limited studies into associations between social determinants and LCAS, including cognitive impairment.^6^

In our prior work in the Tufts Vascular Neurology Registry, we found that higher neighborhood income inequality as measured by the Gini index (a measure of group socioeconomic status variability) was associated with post-stroke functional independence.^7^ A more frequently studied measure of socioeconomic status among patients with stroke is the Area Deprivation Index (ADI). The ADI, reported in deciles, measures how deprived neighborhoods are in comparison to one another.^8,9^ Previous work has also indicated those living in high ADI neighborhoods (high deprivation) have worse initial stroke severity as well as a larger infarct size on initial presentation.^10^ To our knowledge, these group measures have not previously been combined to assess associations in the most vulnerable population (low Gini, high ADI, or homogeneously deprived neighborhoods) with post-stroke outcomes, including LCAS such as cognitive impairment.

Accordingly, we sought in this study to assess potential associations between these neighborhood measures of socioeconomic status and post-stroke cognitive impairment. We hypothesized those patients living in homogeneously deprived neighborhoods would have higher likelihood of cognitive impairment diagnosed in the first year after discharge for acute ischemic strokes (AIS).

## Methods

This analysis has IRB approval through the Tufts Vascular Neurology Registry IRB protocol #12151.

### Study Population

This was a retrospective cohort study of patients aged 18 years and older hospitalized at Tufts Medical Center with AIS between 1/1/2018 - 12/31/2022 with subsequent follow-up in the Stroke Clinic where clinicians performed active surveillance for LCAS symptoms between 0-12 months (n = 1133). Patients were excluded if they had repeat hospitalizations for AIS in this 12-month follow-up period. Active surveillance utilized a semi-structured interview performed at the outpatient appointments and was recorded by the use of a specific note template. In a prior study of this active surveillance strategy, LCAS were detected throughout the first 12 months after stroke, in some cases with initial detection later in that time frame (between 3-12 months as opposed to the first three months).^3^ Accordingly, in this analysis, we only included patients who attended at least one follow-up appointment in either the first three months or the 3-12 month period to minimize effects of loss to follow-up on the results (n = 498 patients without follow up data). Since ADI was measured on a state-level scale, only patients with a Massachusetts ZIP code at the time of hospitalization were included. Additionally, only those residing in a neighborhood with an assessed ADI were included (n = 98 patients without ZIP code data). Patients were excluded if they had incomplete covariate data (n = 25). Final study sample size was n = 512 patients (*Figure 1)*.

**Figure 1.**
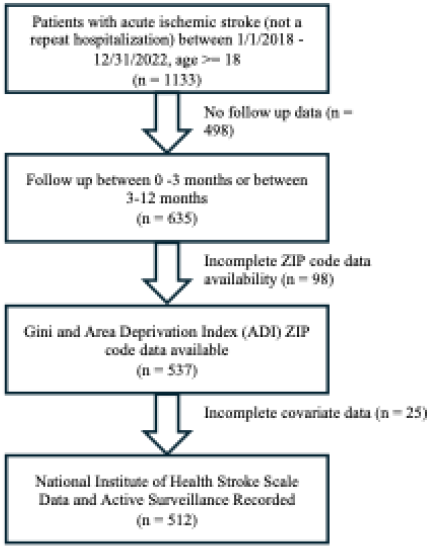
Flow diagram of how the sample size was constructed.

### Predictors and Outcomes

Using the electronic medical record, study variables were collected including age at the time of hospitalization (18-50, 51+), self-reported race (Asian, Black, White, other [or not indicated in the medical record]), self-reported ethnicity (Hispanic, non-Hispanic), sex (male, female), past medical history (PMH), stroke severity (NIHSS on admission), acute revascularization treatments (intravenous thrombolysis, IVT, and endovascular therapy, EVT), active surveillance of LCAS status as determined by use of specific note template, ZIP code at the time of hospitalization, ADI using the address at the time of hospitalization, and cognitive impairment detection at appointments 3 and 12 months after hospitalization. The patient’s ZIP code at the time of hospitalization was used to obtain their neighborhood poverty rate as recorded by the US Census published in 2021 which was dichotomized at the sample median (9.8%).^11^

The primary predictor variables were the Gini and ADI. Both were recorded using patient registration data most recent to their hospitalization. Neighborhood Gini values by ZIP code were recorded using US Census data freely available online. Gini values are published by zip code in 5-year periods by the Census as a part of the American Community Survey. We used the report published in 2021, the most recent at the time of data collection.^12^ ADI decile values by address were obtained utilizing the Neighborhood Atlas, a free online resource. The 2021 report was utilized as the most current version.^8,9^ Patients were split at the Massachusetts state median Gini value (0.4302) and the 50^th^ percentile Massachusetts ADI. This was done to stratify the patients as living in either low or high Gini and ADI areas in context of the state. Patients living in both a low Gini and a high ADI neighborhood were identified as those living in a homogeneously deprived area.

Cognitive impairment was evaluated at visits within three months and between three to twelve months after discharge. Due to limited data, pre-stroke cognitive impairment was not included. Cognitive impairment was defined as detection of cognitive impairment at either of these visits. The use of active surveillance was recorded indicating semi-structured interview questions as outlined in previous studies.^3^ This was included as a covariate as our prior studies have shown that active surveillance is strongly associated with increased detection of LCAS.^3^ Symptoms were recorded as documented in clinic notes from the electronic medical record. If any documentation was unclear, then a senior study investigator (LYL) would review for a final adjudication.

### Statistical Analysis

Statistical analysis was conducted using Stata v18.5, College Station, Texas. Univariate chi-square and T-tests were conducted comparing the variables of interest between Gini, ADI, and the combination Gini-ADI groups.

The primary logistic regression model was constructed to evaluate the likelihood of cognitive impairment. Cognitive impairment was included as a binary variable determining if it was documented at either the 0-3 or 3-12 months follow-up. The model adjusted for age, race, ethnicity, sex, stroke severity, IVT, ZIP code poverty rate, and active surveillance. We dichotomized the poverty rate at the median. We dichotomized the NIHSS at less than or equal to 9 or greater than or equal to 10. Prior research suggested this split to reflect clinical outcomes as patients who have a NIHSS 10 or greater have an increased risk of poor outcomes after stroke.^13^ EVT was excluded due to collinearity in our data with both NIHSS, which is a significant determinant in the usage of EVT (OR: 24.85, 95% CI: 13.88 – 44.50),^14^ and with IVT (OR: 4.24, 95% CI: 2.69 – 6.69). The model was constructed by stepwise addition of variables selected by clinical expertise and prior literature. Likelihood of cognitive impairment was compared in relation to Gini, ADI, and the combination Gini-ADI groups. Variance inflation factor for the models, and all variables had values below 2.

Two sensitivity analyses were conducted. First, we stratified patients based on race and ethnicity. We did a sub-analysis of the 221 not identified as White and non-Hispanic in the study population. Then, we conducted an analysis of the 291 White non-Hispanic patients. This was done to evaluate if racial and ethnic differences may be an effect modifier. We considered evaluating each race and ethnicity separately, however, these subgroups were too small for effective analysis. In the second sensitivity analysis we included the addition of PMH variables that were found to be significantly different across groups in the univariate analysis. These variables were atrial fibrillation and diabetes.

## Results

### Baseline Characteristics

We included 512 patients in the analysis, of whom 206 (40.2%) patients lived in an area with a Gini value below the state median, 281 (54.9%) patients lived in an area with an ADI above the state median, and 140 (27.3%) patients lived in an area with both a low Gini and a high ADI value. The mean NIHSS on presentation was 7. Most patients included had less severe strokes as 70.1% had NIHSS 9 or less and 29.9% had NIHSS 10 or greater. Active surveillance was evenly distributed with 51.8% with active surveillance and 48.2% of patients without. The most common PMH diagnosis was hypertension with 352 (68.8%) patients being identified. Cognitive impairment after stroke affected 154 (30.1%) patients (*Table 1)*.

**Table 1.** Univariate comparison of patient characteristics.

|  | High Gini | Low Gini | P | Low ADI | High ADI | P | Not High ADI and low Gini | High ADI and low Gini | P | Total |
| --- | --- | --- | --- | --- | --- | --- | --- | --- | --- | --- |
| N (%) | 306 (59.8%) | 206 (40.2%) |  | 231 (45.1%) | 281 (54.9%) |  | 372 (72.7%) | 140 (27.3%) |  | 512 (100.0%) |
| Female (%) | 128 (41.8%) | 87 (42.2%) | 0.928 | 90 (39.0%) | 125 (44.5%) | 0.208 | 154 (41.4%) | 61 (43.6%) | 0.657 | 215 (42.0%) |
| Male (%) | 178 (58.2%) | 119 (57.8%) |  | 141 (61.0%) | 156 (55.5%) |  | 218 (58.6%) | 79 (56.4%) |  | 297 (58.0%) |
| Non-Hispanic White (%) | 128 (41.8%) | 163 (79.1%) | <0.001 | 123 (53.2%) | 168 (59.8%) | 0.004 | 177 (47.6%) | 114 (81.4%) | <0.001 | 291 (56.8%) |
| Non-Hispanic Asian (%) | 106 (34.6%) | 17 (8.3%) |  | 73 (31.6%) | 50 (17.8%) |  | 115 (30.9%) | 8 (5.7%) |  | 123 (24.0%) |
| Non-Hispanic Black (%) | 47 (15.4%) | 12 (5.8%) |  | 20 (8.7%) | 39 (13.9%) |  | 52 (14.0%) | 7 (5.0%) |  | 59 (11.5%) |
| Hispanic (%) | 19 (6.2%) | 7 (3.4%) |  | 9 (3.9%) | 17 (6.0%) |  | 21 (5.6%) | 5 (3.6%) |  | 26 (5.1%) |
| Other Race or Ethnicity (%) | 6 (2.0%) | 7 (3.4%) |  | 6 (2.6%) | 7 (2.5%) |  | 7 (1.9%) | 6 (4.3%) |  | 13 (2.5%) |
| Age 18-50 (%) | 41 (13.4%) | 33 (16.0%) | 0.408 | 38 (16.5%) | 36 (12.8%) | 0.244 | 53 (14.2%) | 21 (15.0%) | 0.829 | 74 (14.5%) |
| Age 51+ (%) | 265 (86.6%) | 173 (84.0%) |  | 193 (83.5%) | 245 (87.2%) |  | 319 (85.8%) | 119 (85.0%) |  | 438 (85.5%) |
| High Poverty Neighborhood (%) | 227 (74.2%) | 24 (11.7%) | <0.001 | 108 (46.8%) | 143 (50.9%) | 0.352 | 230 (61.8%) | 21 (15.0%) | <0.001 | 251 (49.0%) |
| High NIHSS >= 10 (%) | 72 (23.5%) | 81 (39.3%) | <0.001 | 51 (22.1%) | 102 (36.3%) | <0.001 | 93 (25.0%) | 60 (42.9%) | <0.001 | 153 (29.9%) |
| Active Surveillance (%) | 149 (48.7%) | 116 (56.3%) | 0.091 | 124 (53.7%) | 141 (50.2%) | 0.430 | 191 (51.3%) | 74 (52.9%) | 0.760 | 265 (51.8%) |
| Cognitive Impairment (%) | 86 (28.1%) | 68 (33.0%) | 0.235 | 70 (30.3%) | 84 (29.9%) | 0.920 | 105 (28.2%) | 49 (35.0%) | 0.136 | 154 (30.1%) |
| Atrial Fibrillation (%) | 59 (19.3%) | 58 (28.2%) | 0.019 | 44 (19.0%) | 73 (26.0%) | 0.063 | 74 (19.9%) | 43 (30.7%) | 0.009 | 117 (22.9%) |
| Diabetes (%) | 98 (32.0%) | 48 (23.3%) | 0.032 | 66 (28.6%) | 80 (28.5%) | 0.980 | 115 (30.9%) | 31 (22.1%) | 0.050 | 146 (28.5%) |
| Hypertension (%) | 210 (68.6%) | 142 (68.9%) | 0.942 | 160 (69.3%) | 192 (68.3%) | 0.820 | 260 (69.9%) | 92 (65.7%) | 0.363 | 352 (68.8%) |
| Hyperlipidemia (%) | 143 (46.7%) | 96 (46.6%) | 0.977 | 108 (46.8%) | 131 (46.6%) | 0.976 | 180 (48.4%) | 59 (42.1%) | 0.207 | 239 (46.7%) |

Demographic variables varied by Gini, ADI, and Gini-ADI groups as seen in Table 1. Notably, Chi-square analyses indicated race and ethnicity varied significantly between Gini (p < 0.001), ADI (p = 0.004), and ADI-Gini groups (p < 0.001). There were no significant differences by age (*Table 1*).

Differences between PMH variables were seen. More patients in the low Gini and the high ADI – low Gini groups had atrial fibrillation (p = 0.019, p = 0.009, respectively). The low Gini and high ADI – low Gini groups also were less likely to have diabetes (p = 0.032, p = 0.050, respectively) (*Table 1*).

NIHSS on admission was high for those in low Gini neighborhoods, with 39.3% of patients living in low Gini areas having an initial NIHSS 10 or higher while only 23.5% of patients from high Gini areas did (p < 0.001). The high ADI group presented with a high NIHSS 36.3% of the time while the low ADI group only did so 22.1% of the time (p < 0.001). Initial NIHSS was high in 42.9% of patients in the high ADI – low Gini group while it was high in only 25.0% of the rest of the sample (p < 0.001) (*Table 1*) (*Figure 2)*.

**Figure 2.**
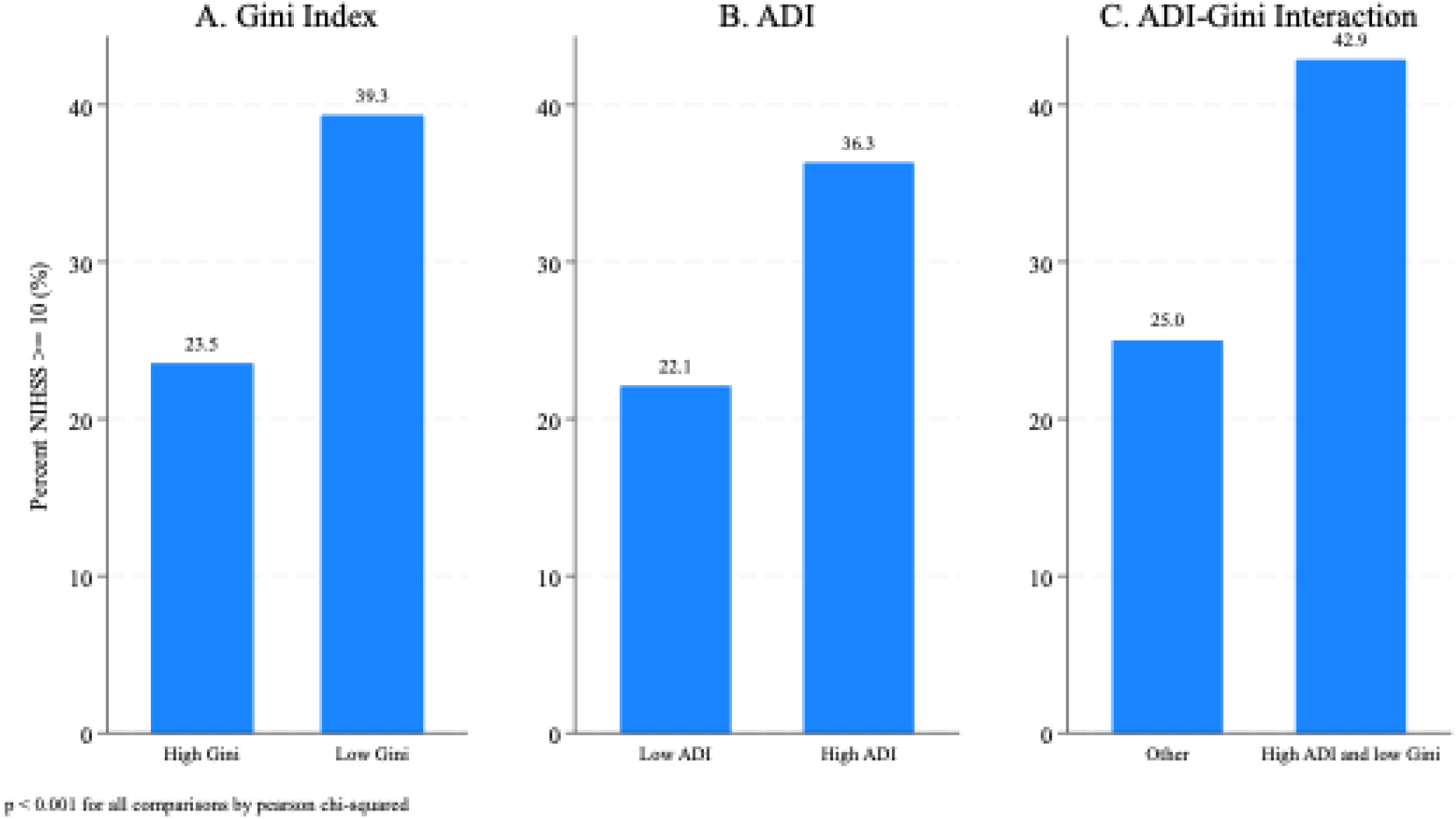
Initial NIH Stroke Scale by Socioeconomic Status. Bar charts indicating the percentage of patients presenting with a severe stroke (initial NIH Stroke Scale >= 10) by ZIP code socioeconomic status groups. A: Gini index measuring ZIP code inequality (higher Gini corresponds to more inequality). B: Area Deprivation Index (ADI) measuring ZIP code disadvantage (higher ADI corresponds to more disadvantage). C: High ADI and Low Gini combination measuring both inequality and disadvantage (high ADI and low Gini correspond to homogeneously disadvantaged).

### Socioeconomic disparities in late complications

Likelihood of cognitive impairment was compared across levels of ADI and Gini. Cognitive impairment was not associated with either ADI or Gini in the univariate analysis (p = 0.920, p = 0.235, respectively). The high ADI – low Gini group had a 35.0% likelihood of cognitive impairment compared to only 28.2% in the remainder of the study population (p = 0.136) (*Table 1*).

In the primary model adjusting for sex, race and ethnicity, poverty, active surveillance, age, NIHSS, and IVT, low Gini was not significantly associated with cognitive impairment (OR: 1.74, 95% CI: 0.98 – 3.07) (*Table 2*). The adjusted analysis by ADI alone was not significantly associated with cognitive impairment (OR: 1.03, 95% CI: 0.67 – 1.57) (*Table 2*). However, likelihood of cognitive impairment was found to be elevated in the high ADI – low Gini group compared to the remainder of participants (all other combinations of ADI and Gini) (OR: 1.82, 95% CI: 1.08 - 3.06). In this model, active surveillance was also significantly positively associated with detection of cognitive impairment (OR: 6.92, 95% CI: 4.33 - 11.06) (*Table 2*).

**Table 2.** Logistic Regressions Comparing Cognitive Impairment by ADI, Gini and by the Combination of High ADI and Low Gini.

|  | Analysis by Low Gini |  | Analysis by High ADI |  | Analysis by High ADI and Low Gini |  |
| --- | --- | --- | --- | --- | --- | --- |
| Cognitive Impairment | Odds Ratio | 95% Confidence Interval | Odds Ratio | 95% Confidence Interval | Odds Ratio | 95% Confidence Interval |
| Male | 0.89 | 0.58 - 1.36 | 0.88 | 0.57 - 1.34 | 0.91 | 0.59 - 1.39 |
| Non-Hispanic White | 1.00 |  | 1.00 |  | 1.00 |  |
| Non-Hispanic Asian | 0.99 | 0.55 - 1.76 | 0.93 | 0.52 - 1.65 | 1.01 | 0.56 - 1.80 |
| Non-Hispanic Black | 0.81 | 0.40 - 1.64 | 0.78 | 0.39 - 1.60 | 0.83 | 0.41 - 1.70 |
| Hispanic | 0.82 | 0.29 - 2.37 | 0.82 | 0.28 - 2.36 | 0.85 | 0.29 - 2.44 |
| Other Race or Ethnicity | 1.53 | 0.44 - 5.28 | 1.47 | 0.43 - 5.06 | 1.39 | 0.39 - 4.86 |
| Age (51+) | 1.36 | 0.76 - 2.42 | 1.37 | 0.77 - 2.45 | 1.37 | 0.77 - 2.46 |
| Active Surveillance | 6.69 | 4.19 - 10.67 | 6.76 | 4.25 - 10.77 | 6.92 | 4.33 - 11.06 |
| High NIHSS $\geq 10$ | 0.90 | 0.56 - 1.45 | 0.91 | 0.57 - 1.47 | 0.89 | 0.55 - 1.42 |
| IVT | 1.05 | 0.66 - 1.69 | 1.09 | 0.68 - 1.75 | 1.05 | 0.65 - 1.68 |
| High Poverty | 2.01 | 1.11 - 3.64 | 1.44 | 0.89 - 2.34 | 1.76 | 1.05 - 2.94 |
| Low Gini | 1.74 | 0.98 - 3.07 |  |  |  |  |
| High ADI |  |  | 1.03 | 0.67 - 1.57 |  |  |
| High ADI and Low Gini |  |  |  |  | 1.82 | 1.08 - 3.06 |

The first sensitivity analysis stratified non-Hispanic White patients and all patients that were not non-Hispanic White. Among 291 non-Hispanic White patients, 114 (39.2%) lived in a high ADI and low Gini neighborhood. Among the 221 remaining patients of any other racial or ethnic group, 26 (11.8%) lived in a high ADI and low Gini neighborhood. For these patients that were not classified as non-Hispanic White, the effect was amplified. The high ADI – low Gini group had almost triple the odds of having cognitive impairment detected (OR: 3.38, 95% CI: 1.11 – 10.30) (*Table 3*). The effect was not seen when conducting the analysis with the non-Hispanic White patients (OR: 1.49, 95% CI: 0.82 - 2.69) (*Table 3*).

**Table 3.** Logistic Regression Sensitivity Analyses Comparing Cognitive Impairment by Neighborhood Gini and ADI When Stratifying by Race and Ethnicity and When Including Past Medical History Covariates.

|  | All patients Not Non-Hispanic White (n = 221) |  | Non-Hispanic White Patients (n = 291) |  | All Patients and PMH Included (n = 512) |  |
| --- | --- | --- | --- | --- | --- | --- |
| Cognitive Impairment | Odds Ratio | 95% Confidence Interval | Odds Ratio | 95% Confidence Interval | Odds Ratio | 95% Confidence Interval |
| Male | 0.71 | 0.36 - 1.40 | 1.10 | 0.62 - 1.93 | 0.91 | 0.60 - 1.40 |
| Non-Hispanic White |  |  |  |  | 1.00 |  |
| Non-Hispanic Asian | 1.00 |  |  |  | 1.00 | 0.56 - 1.79 |
| Non-Hispanic Black | 0.84 | 0.39 - 1.81 |  |  | 0.84 | 0.41 - 1.73 |
| Hispanic | 0.85 | 0.28 - 2.62 |  |  | 0.85 | 0.30 - 2.47 |
| Other Race or Ethnicity | 1.42 | 0.31 - 6.41 |  |  | 1.30 | 0.37 – 4.62 |
| Age (51+) | 2.07 | 0.81 – 5.26 | 1.05 | 0.50 – 2.22 | 1.28 | 0.70 – 2.31 |
| Active Surveillance | 8.77 | 4.15 - 18.54 | 5.63 | 3.06 – 10.34 | 6.97 | 4.35 – 11.16 |
| High NIHSS >= 10 | 0.81 | 0.35 - 1.89 | 0.92 | 0.51 - 1.63 | 0.83 | 0.51 - 1.34 |
| IVT | 0.91 | 0.38 - 2.16 | 1.12 | 0.63 - 1.98 | 1.07 | 0.67 - 1.73 |
| Atrial Fibrillation |  |  |  |  | 1.41 | 0.85 - 2.36 |
| Diabetes |  |  |  |  | 1.03 | 0.64 - 1.63 |
| High Poverty | 2.17 | 0.85 – 5.55 | 1.50 | 0.79 – 2.82 | 1.79 | 1.06 – 3.01 |
| High ADI and Low Gini | 3.38 | 1.11 - 10.30 | 1.49 | 0.82 - 2.69 | 1.79 | 1.06 – 3.02 |

The second sensitivity analysis included the additional variables atrial fibrillation and diabetes as these were significantly different between socioeconomic groups in the univariate analysis. The high ADI – low Gini group was significantly associated with cognitive impairment (OR: 1.79, 95% CI: 1.06 – 3.02) (*Table 3*). Neither atrial fibrillation nor diabetes diagnosis was significantly associated with cognitive impairment in this model. The analyses by Gini or ADI alone were not significant.

## Discussion

To our knowledge, this is the first study evaluating the combination of ADI and Gini and its associations with post-stroke cognitive impairment at a center leveraging active surveillance for LCAS. In adjusted analyses, we found that patients living in communities that are homogeneously deprived – high ADI and low Gini – were more likely to have cognitive impairment detected within one year after stroke. No associations of Gini or ADI alone with post-stroke cognitive impairment were found in either univariate or multivariable analyses. We further demonstrated that this association is only present for non-White patients. This study underscores the potential influence of neighborhood socioeconomic factors on clinically meaningful post-stroke outcomes.

This study combined ADI and Gini to provide a multidimensional understanding of neighborhood socioeconomic status and its potential effects for patients with acute stroke. ADI is increasingly commonly studied to assess health care disparities in stroke and other medical conditions, but it provides limited understanding of resource variance within a neighborhood.^10,15^ By contrast, Gini offers unique insights on variability but does not account for the overall resource availability of a neighborhood. Both ADI and Gini alone were not associated with post-stroke cognitive impairment in our study. One possible reason why we see these findings is due to a homogenous lack of resources in the neighborhood which magnifies disadvantage. Another possibility is that heterogeneously deprived areas are difficult to study at the neighborhood level statistically. Since our measure is at the neighborhood level and the neighborhood has large amounts of inequality, both those with more or less access to resources will be categorized the same way, diminishing any effect. Using ADI and Gini in conjunction allowed us to have a more nuanced analysis of the socioeconomic effects of stroke outcomes. Further studies leveraging this combined measure for the analysis of other health outcomes are warranted.

Our study had several strengths. First, about half of the patients in this study underwent routine active surveillance of LCAS, likely enhancing detection of post-stroke cognitive impairment and providing a more accurate assessment of the true frequency of this complication during the first year after ischemic stroke.^3^ Second, the study took place at a tertiary care, academic Comprehensive Stroke Center with a large catchment area including the Eastern half of Massachusetts. The catchment area included a wide variety of neighborhoods with diverse demographics and socioeconomic status. Boston has one of the highest Gini indices of any city in the US.^16^ Regarding limitations, this study did not collect information on pre-stroke cognitive impairment or dementia, so it is not known if risks of these conditions differed across levels of ADI and Gini. Additionally, cognitive impairment was identified with a pragmatic screening tool but was not uniformly confirmed through a validated diagnostic tool (e.g. Montreal Cognitive Assessment). We also categorized age into ≤50/≥51 so we cannot comment on variability within age ranges.

## Conclusions

Our study indicates those living in homogeneously deprived neighborhoods are more likely to have detection of post-stroke cognitive impairment. This study indicates a need for targeted interventions in communities most at risk. Future studies should evaluate a more granular examination of these effects and the causes behind them. Additionally, this study highlights the use of combined socioeconomic status measures in stroke outcome studies. Future studies should utilize this approach in evaluation of complex socioeconomic effects that are not explained by one variable alone.

## Author Contributions

Matthew Siegel: Conceptualization, Methodology, Investigation, Formal Analysis, Writing – Original Draft. Laura Corlin: Methodology, Writing – Review and Editing. James Miller: Investigation, Writing – Review and Editing. Kathryn Cote: Investigation, Writing – Review and Editing. Lester Leung: Conceptualization, Methodology, Writing – Review and Editing, Supervision.

## Data Availability

Data available on request

## References

1. Jones AA, Zhou LW, Castro N, et al. Novel Network Analysis of County-and Individual-Level Factors Associated With Functional Outcomes After Stroke. Stroke. 2025;56(5):1180–1190. doi:10.1161/STROKEAHA.124.048336

2. Taghdiri F, Vyas MV, Kapral MK, et al. Association of Neighborhood Deprivation With Thrombolysis and Thrombectomy for Acute Stroke in a Health System With Universal Access. Neurology. 2023;101(22):e2215–e2222. doi:10.1212/WNL.0000000000207924

3. Gans SD, Michaels E, Thaler DE, Leung LY. Detection of symptoms of late complications after stroke in young survivors with active surveillance versus usual care. Disabil Rehabil. 2022;44(15):4023–4028. doi:10.1080/09638288.2021.1883749

4. Sun JH, Tan L, Yu JT. Post-stroke cognitive impairment: epidemiology, mechanisms and management. Ann Transl Med. 2014;2(8):80. doi:10.3978/j.issn.2305-5839.2014.08.05

5. Chohan SA, Venkatesh PK, How CH. Long-term complications of stroke and secondary prevention: an overview for primary care physicians. Singapore Med J. 2019;60(12):616. doi:10.11622/smedj.2019158

6. Tian J, Wang Y, Guo L, Li S. Association of Income with Post-Stroke Cognition and the Underlying Neuroanatomical Mechanism. Brain Sci. 2023;13(2):2. doi:10.3390/brainsci13020363

7. Coté KE, Pudlo ME, Jost-Price E, Leung LY. Neighborhood income inequality associated with functional independence after ischemic stroke: a cohort study. J Stroke Cerebrovasc Dis. 2025;34(1):108035. doi:10.1016/j.jstrokecerebrovasdis.2024.108035

8. University of Wisconsin School of Medicine and Public Health. 2021 Area Deprivation Index v4.0.1. Accessed January 23, 2024. https://www.neighborhoodatlas.medicine.wisc.edu

9. Kind AJH, Buckingham WR. Making Neighborhood-Disadvantage Metrics Accessible — The Neighborhood Atlas. N Engl J Med. 2018;378(26):2456–2458. doi:10.1056/NEJMp1802313

10. Ghoneem A, Osborne MT, Abohashem S, et al. Association of Socioeconomic Status and Infarct Volume With Functional Outcome in Patients With Ischemic Stroke. JAMA Netw Open. 2022;5(4):e229178. doi:10.1001/jamanetworkopen.2022.9178

11. U.S. Census Bureau. POVERTY STATUS IN THE PAST 12 MONTHS. Accessed February 26, 2023. https://data.census.gov/table/ACSST5Y2021.S1701?g=860XX00US02144

12. US Census Bureau. GINI INDEX OF INCOME INEQUALITY. Accessed February 26, 2023. https://data.census.gov/table/ACSDT5Y2021.B19083?q=B19083:+GINI+INDEX+OF+INCOME+INEQUALITY&g=040XX00US25,25$8600000

13. Qureshi AI, Bhatti IA, Gillani SA, Fakih R, Gomez CR, Kwok CS. Factors and outcomes associated with National Institutes of Health stroke scale scores in acute ischemic stroke patients undergoing thrombectomy in United States. J Stroke Cerebrovasc Dis Off J Natl Stroke Assoc. 2025;34(6):108292. doi:10.1016/j.jstrokecerebrovasdis.2025.108292

14. Powers WJ, Rabinstein AA, Ackerson T, et al. Guidelines for the Early Management of Patients With Acute Ischemic Stroke: 2019 Update to the 2018 Guidelines for the Early Management of Acute Ischemic Stroke: A Guideline for Healthcare Professionals From the American Heart Association/American Stroke Association. Stroke. 2019;50(12):e344–e418. doi:10.1161/STR.0000000000000211

15. Morenz AM, Liao JM, Au DH, Hayes SA. Area-Level Socioeconomic Disadvantage and Health Care Spending: A Systematic Review. JAMA Netw Open. 2024;7(2):e2356121. doi:10.1001/jamanetworkopen.2023.56121

16. Ranking Cities by the New Urban Crisis. Bloomberg.com. August 29, 2019. Accessed November 4, 2024. https://www.bloomberg.com/news/articles/2019-08-29/ranking-cities-by-the-new-urban-crisis

